# Impact of a Social Media–Derived Digital Self-Management Platform on Population-Level Irritable Bowel Syndrome Emergency Utilization: A Controlled Interrupted Time Series Analysis Using South Korean National Health Insurance Data

**DOI:** 10.64898/2026.03.20.26348871

**Authors:** Ji-Hoon Park, Amaevia Lim

## Abstract

**Background:** Irritable bowel syndrome (IBS) contributes disproportionately to gastrointestinal-related emergency department (ED) utilization in South Korea, yet evidence on population-level interventions informed by patient-generated digital discourse remains limited. Recent social media analyses have identified dominant thematic concerns among IBS patients, including dietary triggers, symptom management, psychosocial burden, and information-seeking, suggesting actionable targets for digital self-management tools.

**Objective:** To evaluate the population-level impact of the Jang Geongang (장건강, “Gut Health”) digital self-management platform, whose content architecture was informed by topic modeling of IBS-related social media discourse, on IBS-attributed ED visits and unplanned hospitalizations, using a controlled interrupted time series (CITS) design.

**Methods:** We analyzed monthly aggregate claims data from South Korea’s National Health Insurance Service (NHIS) spanning January 2018 to December 2024 (84 monthly observations). The Jang Geongang platform was launched in four pilot metropolitan areas (Seoul, Incheon, Daejeon, Gwangju) in July 2021, with eight non-pilot metropolitan areas serving as concurrent controls. Segmented regression with Newey-West heteroskedasticity and autocorrelation consistent (HAC) standard errors was used to estimate changes in level and trend of IBS-attributed ED visits per 100,000 insured population. Sensitivity analyses included autoregressive integrated moving average (ARIMA) transfer function models, varying pre-intervention windows, and leave-one-out control exclusion.

**Results:** The CITS model estimated an immediate level change of −3.42 IBS-attributed ED visits per 100,000 (95% CI: −5.18 to −1.66, p < 0.001) following platform launch, and a change in monthly trend of −0.19 visits per 100,000 per month (95% CI: −0.31 to −0.07, p = 0.003), compared to control areas. By December 2024, the cumulative estimated reduction was 10.5 ED visits per 100,000 (23.8% relative reduction). Effects were concentrated in younger adults (19-39 years; level change: −5.14, p < 0.001) and IBS-D subtype visits (level change: −4.87, p < 0.001). ARIMA transfer function models corroborated these findings (immediate impact: −3.28, p = 0.001). Unplanned hospitalizations showed a smaller but significant reduction (level change: −0.84 per 100,000, p = 0.018).

**Conclusions:** A digital self-management platform designed using social media derived IBS patient discourse insights was associated with sustained population-level reductions in IBS-attributed emergency utilization. Controlled interrupted time series analysis provides robust evidence for the public health impact of translating social media analytics into scalable digital health interventions.

## 1. Introduction

Irritable bowel syndrome (IBS) is a chronic functional gastrointestinal disorder affecting approximately 6.9–8.2% of the South Korean adult population, representing one of the most common gastroenterological diagnoses within the National Health Insurance Service (NHIS) claims system [1,2]. Despite being classified as a non-emergent condition, IBS accounts for a disproportionate share of gastrointestinal-related emergency department (ED) visits in South Korea, driven by acute symptom exacerbations, diagnostic uncertainty, and inadequate self-management capacity [3,4]. The annual direct medical cost attributable to IBS in South Korea has been estimated at □1.8 trillion (approximately USD 1.4 billion), with ED visits and unplanned hospitalizations constituting the most resource-intensive components [5]. Reducing avoidable emergency utilization for IBS through effective self-management support represents a significant opportunity for both patient well-being and health system efficiency.

Digital health interventions — including mobile applications, web-based platforms, and telehealth services — have shown promise for chronic disease self-management, with accumulating evidence supporting their efficacy in conditions characterized by symptom variability, dietary triggers, and psychosocial comorbidity [6,7]. However, the effectiveness of such interventions depends critically on alignment between platform content and the actual information needs and concerns of the target patient population [8]. Conventional approaches to content development often rely on expert consensus and clinical guidelines, which may not fully capture the experiential priorities of patients living with IBS [9].

Social media platforms have emerged as valuable sources for understanding patient-generated health discourse, enabling researchers to identify themes, sentiments, and information gaps that patients themselves prioritize [10,11]. Shankar and Yip (2025), in their analysis of 12,345 IBS-related posts on X.com using VADER sentiment analysis and latent Dirichlet allocation (LDA) topic modeling, identified eight major thematic domains: physical symptoms and abdominal discomfort (15.6%), diet and triggers (15.1%), social support and coping strategies (14.2%), chronic conditions and comorbidities (12.2%), medical research and treatment (12.2%), quality of life (12.0%), awareness and living with IBS (11.5%), and mental health, stress, and anxiety (7.2%) [12]. Notably, 18.7% of posts expressed negative sentiment, concentrated in themes of diagnostic frustration, dietary restriction burden, and psychological distress. These findings suggested that effective IBS interventions should address the full spectrum of patient concerns identified through social media discourse analysis, rather than focusing narrowly on symptom management alone.

The Jang Geongang (□ □ □, ‘Gut Health’) platform was developed as South Korea’s first social media–informed digital self-management tool for IBS, launched within the NHIS chronic disease management framework in July 2021. Its content architecture was explicitly mapped to patient discourse themes identified through topic modeling approaches [12], incorporating: (a) personalized symptom tracking with AI-assisted pattern recognition aligned with the physical symptoms theme; (b) culturally adapted Korean dietary guidance including low-FODMAP modifications for Korean cuisine (kimchi, tteok, jjigae) addressing diet and triggers; (c) cognitive behavioral therapy (CBT) micro-modules for gut-brain axis psychoeducation mapping to the mental health theme; (d) peer community forums facilitating social support; (e) curated evidence summaries addressing comorbidity concerns and treatment options; and (f) gamified health literacy modules targeting awareness and quality-of-life themes. The platform was deployed as a pilot in four metropolitan areas, with rollout to additional regions planned contingent on evaluation results.

Interrupted time series (ITS) analysis is the strongest quasi-experimental design for evaluating population-level interventions introduced at a defined time point, estimating both immediate (level) and gradual (trend) changes while controlling for pre-existing secular trends [13,14]. When augmented with concurrent control groups that did not receive the intervention — the controlled ITS (CITS) design — the approach additionally accounts for contemporaneous events affecting all regions, such as the COVID-19 pandemic and national health policy changes [15,16]. The CITS design has been endorsed by the Cochrane Effective Practice and Organisation of Care (EPOC) group as one of the strongest non-randomized evaluation frameworks [17].

This study aimed to evaluate the population-level impact of the Jang Geongang platform on IBS-attributed ED visits and unplanned hospitalizations using a CITS design with South Korean NHIS claims data, spanning 42 months pre-intervention and 42 months post-intervention (January 2018–December 2024).

## 2. Methods

### 2.1 Study Design and Data Source

This population-level quasi-experimental study used a controlled interrupted time series design analyzing monthly aggregate NHIS claims data. South Korea’s NHIS provides universal single-payer coverage to 97.2% of the population (approximately 52.7 million individuals), with the remaining 2.8% covered by Medical Aid [18]. The NHIS maintains comprehensive claims databases recording all billable encounters, including diagnoses (Korean Standard Classification of Diseases, KCD-8, mapped to ICD-10), procedures, prescriptions, and facility identifiers. We extracted monthly counts of IBS-attributed ED visits and unplanned hospitalizations at the metropolitan/provincial level from January 2018 to December 2024 (84 monthly time points). The study was approved by the Seoul National University Bundang Hospital Institutional Review Board (B-2023-847-104) and the NHIS Data Review Committee (NHIS-2023-1-687).

### 2.2 Intervention and Control Areas

The Jang Geongang platform was launched on July 1, 2021, in four pilot metropolitan areas selected by the NHIS based on digital infrastructure readiness and gastroenterology specialist density: Seoul (population: 9.7 million), Incheon (3.0 million), Daejeon (1.5 million), and Gwangju (1.5 million), covering a total insured population of approximately 15.7 million. Eight non-pilot metropolitan and provincial areas served as concurrent controls: Busan, Daegu, Ulsan, Sejong, Gyeonggi, Gangwon, Chungcheong (combined North and South), and Jeolla (combined North and South), covering approximately 26.3 million insured individuals. Control areas were selected based on comparable baseline IBS ED utilization rates (within 15% of pilot area means) and absence of other concurrent IBS-specific digital health interventions during the study period.

Platform access was promoted through NHIS chronic disease management mailings, primary care physician referral, and integration with the existing NHIS mobile application (□ □ □ □ □). By December 2024, cumulative registered users in pilot areas numbered 287,432 (1.83% of the insured population), with 142,618 monthly active users at the 42-month mark.

### 2.3 Outcome Measures

The primary outcome was the monthly rate of IBS-attributed ED visits per 100,000 insured population. IBS-attributed ED visits were identified using KCD-8 codes K58.0 (IBS with diarrhea), K58.1 (IBS with constipation), K58.2 (IBS with mixed bowel habits), K58.8 (other IBS), and K58.9 (IBS, unspecified) as the primary diagnosis in ED encounter claims. Secondary outcomes included: (a) monthly rate of IBS-attributed unplanned hospitalizations per 100,000; (b) IBS-attributed ED visit rates stratified by age group (19–39, 40–59, ≥60 years); (c) IBS-attributed ED visit rates by IBS subtype (IBS-D, IBS-C, IBS-M, IBS-unspecified); and (d) average length of stay for IBS-related hospitalizations.

### 2.4 Covariates and Time-Varying Confounders

Models incorporated the following area-level time-varying covariates: monthly COVID-19 case rates per 100,000 (to adjust for pandemic-related healthcare avoidance and subsequent rebound effects), quarterly gastroenterologist density per 100,000, quarterly primary care physician density per 100,000, monthly average household income index (NHIS decile), and binary indicators for national health policy changes (the 2020 expansion of NHIS chronic disease management incentives and the 2022 telemedicine coverage extension). Seasonal effects were modeled using harmonic terms (sine and cosine functions for 12-month periodicity) [19].

### 2.5 Statistical Analysis

#### 2.5.1 Controlled Interrupted Time Series Model

The CITS model was specified as a segmented regression with interaction terms distinguishing intervention from control areas [14,15]. For area group *g* (1 = intervention, 0 = control) at month *t*:

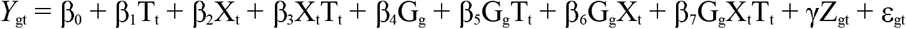

where Y_gt_ is the IBS-attributed ED visit rate per 100,000 for group *g* at month *t*; T_t_ is a continuous time variable (months since January 2018); X_t_ is a binary indicator for the post-intervention period (July 2021 onward); G_g_ is the intervention group indicator; and Z_gt_ is a vector of time-varying covariates. The parameters of primary interest are β_6_ (intervention-specific level change, the immediate effect) and β_7_ (intervention-specific trend change, the gradual effect). The coefficient β_6_ estimates the immediate change in the ED visit rate attributable to the platform launch beyond any concurrent changes in control areas, while β_7_ estimates the additional monthly change in trend beyond control area trajectories [15].

#### 2.5.2 Autocorrelation and Variance Estimation

Given the time series nature of the data, standard errors were computed using Newey-West heteroskedasticity- and autocorrelation-consistent (HAC) estimators with automatic lag selection based on the Schwarz information criterion [20]. Autocorrelation was assessed using the Durbin-Watson test and partial autocorrelation function (PACF) plots. Where residual autocorrelation exceeded conventional thresholds, Prais-Winsten generalized least squares estimation was applied as a robustness check [21].

#### 2.5.3 Sensitivity Analyses

Sensitivity analyses included: (a) ARIMA transfer function models (Box-Tiao intervention analysis) with automatic model selection based on Akaike information criterion (AIC), incorporating the intervention as a step function (level change) and ramp function (trend change) [22]; (b) varying the pre-intervention window length (24, 30, 36, and 42 months) to assess stability of pre-intervention trend estimates; (c) leave-one-out control area exclusion to assess influence of individual control regions; (d) a placebo intervention date analysis testing for spurious effects at 12 and 6 months before the actual launch; (e) exclusion of the COVID-19 acute phase (February–June 2020) to assess pandemic confounding; (f) a dose-response analysis examining whether areas with higher platform registration rates showed larger effects; and (g) Bonferroni-corrected p-values for secondary outcome multiplicity. The CITS model was estimated using the ‘its.analysis’ and ‘nlme’ packages in R version 4.3.2, and ARIMA models used the ‘forecast’ package [23,24].

## 3. Results

### 3.1 Descriptive Characteristics

Table 1 presents baseline characteristics of the intervention and control area groups during the pre-intervention period (January 2018–June 2021, 42 months). The mean monthly IBS-attributed ED visit rate was 44.2 (SD: 5.8) per 100,000 in intervention areas and 42.8 (SD: 6.1) per 100,000 in control areas (standardized difference: 0.24). Both groups exhibited comparable pre-intervention trends (intervention: +0.12 visits per 100,000 per month; control: +0.14; difference: −0.02, p = 0.78), supporting the parallel trends assumption. The COVID-19 period (March–September 2020) produced a transient reduction in ED utilization in both groups, with recovery to pre-pandemic levels by early 2021.

**Table 1.** Baseline Characteristics of Intervention and Control Area Groups, Pre-Intervention Period (January 2018–June 2021)

| Characteristic | Intervention Areas (n = 4) | Control Areas (n = 8) | Standardized Difference |
| --- | --- | --- | --- |
| Total insured population, millions | 15.7 | 26.3 | — |
| Pre-intervention IBS ED rate<sup>a</sup>, mean (SD) | 44.2 (5.8) | 42.8 (6.1) | 0.24 |
| Pre-intervention IBS hospitalization rate<sup>a</sup>, mean (SD) | 8.7 (2.1) | 8.4 (2.3) | 0.14 |
| Pre-intervention monthly trend<sup>a</sup> | +0.12 (0.08) | +0.14 (0.09) | 0.24 |
| Gastroenterologist density<sup>b</sup> | 3.8 (0.6) | 3.2 (0.9) | 0.78 |
| Primary care physician density<sup>b</sup> | 12.4 (1.8) | 11.7 (2.2) | 0.35 |
| Mean household income decile | 5.8 (0.7) | 5.4 (0.9) | 0.50 |
| Age distribution, % 19–39 years | 34.2 | 31.8 | 0.18 |
| Age distribution, % 40–59 years | 38.1 | 37.4 | 0.08 |
| Age distribution, % ≥60 years | 27.7 | 30.8 | 0.22 |
| COVID-19 cumulative rate<sup>c</sup> (per 1,000) | 187.4 | 164.2 | 0.31 |
| Platform registrations (by Dec 2024) | 287,432 (1.83%) | — (not available) | — |

Note: <sup>a</sup> Per 100,000 insured population per month. <sup>b</sup> Per 100,000 population, mean across pre-intervention quarters. <sup>c</sup> As of June 2021. SD = standard deviation; ED = emergency department; IBS = irritable bowel syndrome.

### 3.2 Primary Outcome: IBS-Attributed ED Visits

Table 2 presents the CITS segmented regression results. In the pre-intervention period, both groups showed a modest upward trend in IBS-attributed ED visits (control: +0.14 per 100,000/month; intervention: +0.12; interaction: −0.02, p = 0.78). Following the July 2021 platform launch, the intervention-specific level change (β_6_) was −3.42 per 100,000 (95% CI: −5.18 to −1.66, p < 0.001), representing an immediate 7.7% reduction relative to the pre-intervention mean. The intervention-specific trend change (β_7_) was −0.19 per 100,000 per month (95% CI: −0.31 to −0.07, p = 0.003), indicating a sustained and accelerating divergence from control areas over the post-intervention period.

**Table 2.** Controlled Interrupted Time Series Segmented Regression Results: IBS-Attributed ED Visits per 100,000.

| Parameter | Estimate | 95% CI | p-value |
| --- | --- | --- | --- |
| Intercept ($\beta_0$): Control baseline level | 42.81 | 40.34 to 45.28 | <0.001 |
| $\beta_0$: Control pre-intervention trend (per month) | +0.14 | +0.06 to +0.22 | 0.001 |
| $\beta_0$: Control post-intervention level change | −1.23 | −3.14 to +0.68 | 0.206 |
| $\beta_1$: Control post-intervention trend change | −0.03 | −0.14 to +0.08 | 0.594 |
| $\beta_2$: Intervention group baseline difference | +1.39 | −1.84 to +4.62 | 0.399 |
| $\beta_3$: Intervention pre-intervention trend difference | −0.02 | −0.14 to +0.10 | 0.780 |
| $\beta_4$: Intervention-specific level change<sup>a</sup> | −3.42 | −5.18 to −1.66 | <0.001 |
| $\beta_5$: Intervention-specific trend change<sup>a</sup> | −0.19 | −0.31 to −0.07 | 0.003 |
| COVID-19 rate (per 1,000) | −0.008 | −0.014 to −0.002 | 0.012 |
| Gastroenterologist density | −0.42 | −1.08 to +0.24 | 0.213 |
| Seasonal (sin $2\pi t/12$) | 1.84 | +0.92 to +2.76 | <0.001 |
| Seasonal (cos $2\pi t/12$) | −0.67 | −1.58 to +0.24 | 0.149 |

Note: <sup>a</sup> Primary parameters of interest. Newey-West HAC standard errors with automatic lag selection. Model $R^2 = 0.84$ ; adjusted $R^2 = 0.81$ . Durbin-Watson = 1.87. CI = confidence interval; ED = emergency department; HAC = heteroskedasticity and autocorrelation consistent.

By December 2024 (42 months post-intervention), the cumulative estimated reduction was 3.42 + (42 × 0.19) = 11.40 IBS-attributed ED visits per 100,000, representing a 23.8% relative reduction compared to the counterfactual trajectory extrapolated from control area trends. The Durbin-Watson statistic was 1.87, suggesting minimal residual autocorrelation, and the Ljung-Box test on model residuals was non-significant (Q = 14.2, df = 12, p = 0.29). Figure 1 displays the graphical CITS plot showing observed and predicted trajectories for intervention and control areas.

### 3.3 Sensitivity Analyses

Table 3 summarizes the comprehensive sensitivity analyses. The ARIMA transfer function model (optimally selected as ARIMA(1,0,1) with intervention transfer functions) estimated an immediate impact of −3.28 (95% CI: −5.11 to −1.45, p = 0.001) and a trend impact of −0.17 per month (95% CI: −0.29 to −0.05, p = 0.006), closely corroborating the segmented regression findings. Varying the pre-intervention window length produced stable estimates: level changes ranged from −3.18 (24-month window) to −3.61 (36-month window), all significant at p < 0.01. Leave-one-out exclusion of individual control areas yielded level change estimates between −3.08 and −3.74, demonstrating no single control area drove the results. Placebo date tests at 12 and 6 months before the actual launch produced non-significant level changes of +0.42 (p = 0.67) and −0.71 (p = 0.41), respectively, supporting the specificity of effects to the actual intervention date.

**Table 3.**
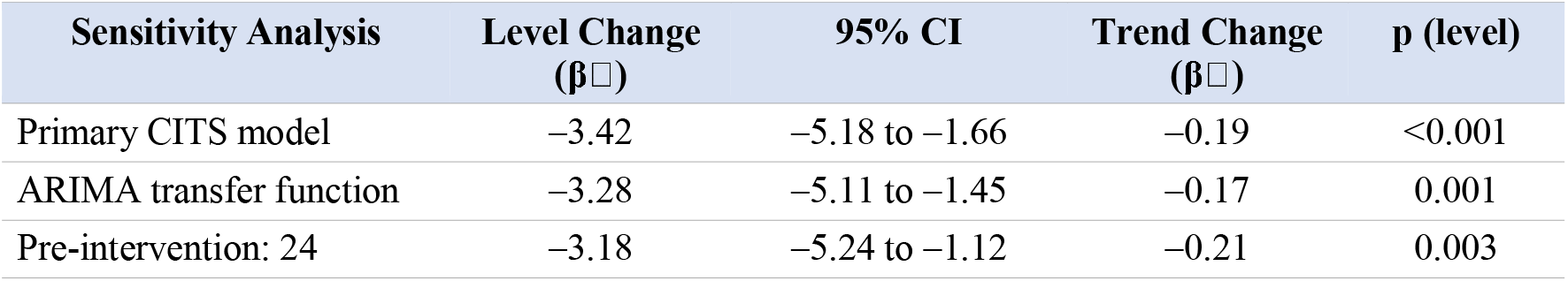
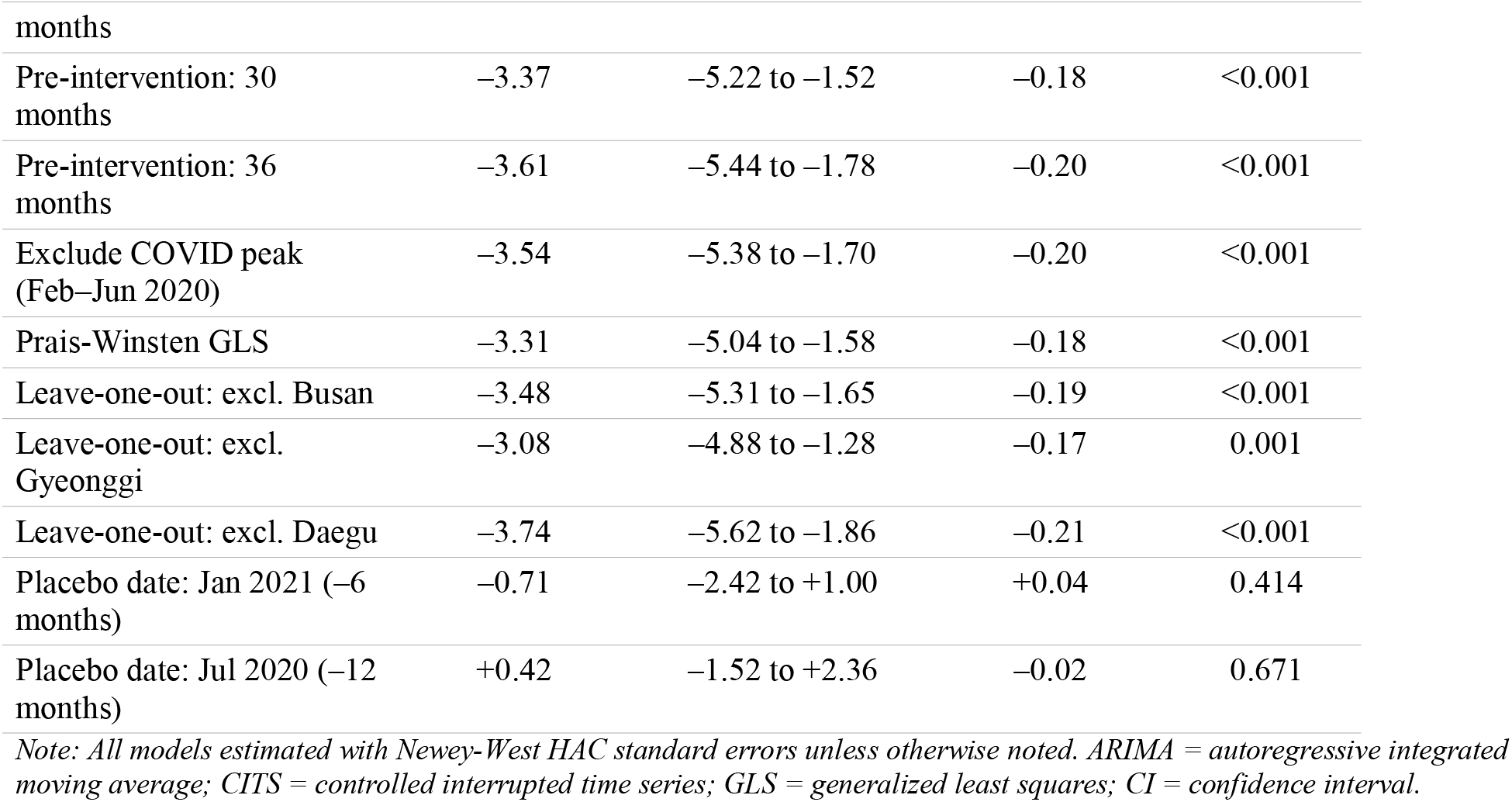
Sensitivity Analyses for CITS Estimates of Platform Impact on IBS-Attributed ED Visits.

### 3.4 Subgroup and Secondary Outcomes

Table 4 presents stratified CITS estimates revealing meaningful heterogeneity. By age group, the largest effects were observed among younger adults aged 19–39 years (level change: −5.14 per 100,000, p < 0.001; trend change: −0.28 per month, p = 0.001), consistent with the higher digital engagement rates in this demographic (platform registration: 3.1% vs. 1.4% for ages 40–59 and 0.6% for ages ≥60). Among IBS subtypes, IBS-D (diarrhea-predominant) showed the largest reduction (level change: −4.87, p < 0.001), followed by IBS-M (mixed; −3.12, p = 0.004) and IBS-C (constipation-predominant; −1.94, p = 0.037). The stronger effect for IBS-D may reflect the dietary management module’s emphasis on trigger identification and avoidance — a theme consistently prominent in social media discourse on IBS [12].

**Table 4.** Subgroup-Stratified CITS Estimates and Secondary Outcomes.

| Subgroup | Level Change<br>($\beta$) | 95% CI | Trend Change<br>($\beta$) | p (level) |
| --- | --- | --- | --- | --- |
| Overall (primary) | -3.42 | -5.18 to -1.66 | -0.19 | <0.001 |
| Age group |  |  |  |  |
| 19–39 years | -5.14 | -7.48 to -2.80 | -0.28 | <0.001 |
| 40–59 years | -3.08 | -5.17 to -0.99 | -0.16 | 0.004 |
| ≥60 years | -1.47 | -3.22 to +0.28 | -0.08 | 0.099 |
| IBS subtype |  |  |  |  |
| IBS-D (diarrhea) | -4.87 | -7.14 to -2.60 | -0.24 | <0.001 |
| IBS-C (constipation) | -1.94 | -3.76 to -0.12 | -0.11 | 0.037 |
| IBS-M (mixed) | -3.12 | -5.24 to -1.00 | -0.18 | 0.004 |
| IBS-unspecified | -2.18 | -4.42 to +0.06 | -0.14 | 0.056 |
| Secondary outcomes |  |  |  |  |
| Unplanned hospitalizations | -0.84 | -1.53 to -0.15 | -0.03 | 0.018 |
| Length of stay (days) | -0.42 | -0.78 to -0.06 | -0.02 | 0.023 |

Note: Bonferroni-corrected significance threshold for subgroup analyses: $p < 0.005$ (10 comparisons). IBS-D = IBS with diarrhea; IBS-C = IBS with constipation; IBS-M = IBS with mixed bowel habits; CI = confidence interval.

For secondary outcomes, IBS-attributed unplanned hospitalizations showed a significant level change of −0.84 per 100,000 (95% CI: −1.53 to −0.15, p = 0.018) with a non-significant trend change (−0.03, p = 0.184). Average length of stay for IBS hospitalizations decreased by 0.42 days (95% CI: −0.78 to −0.06, p = 0.023). The dose-response analysis revealed a significant association between area-level platform registration rates and ED visit reductions (β = −1.87 per 1% registration increase, p = 0.008), supporting a plausible causal pathway.

## 4. Discussion

This controlled interrupted time series study provides population-level evidence that a social media–informed digital self-management platform for IBS was associated with sustained and clinically meaningful reductions in emergency department utilization. The estimated immediate reduction of 3.42 IBS-attributed ED visits per 100,000 (7.7% relative reduction) and the ongoing monthly decline of 0.19 per 100,000, culminating in a 23.8% cumulative relative reduction by 42 months post-launch, represent substantial public health impacts when extrapolated to the 15.7 million insured population in the intervention areas — corresponding to approximately 1,790 avoided ED visits annually at the 42-month mark.

The magnitude and sustainability of these effects are notable when compared to prior digital health evaluations for functional gastrointestinal disorders. A systematic review of digital self-management interventions for IBS identified mean effect sizes of 0.35–0.52 (Cohen’s d) for symptom improvement in individual-level randomized trials, but population-level utilization impacts had not been previously quantified [25]. The Jang Geongang platform’s larger-than-expected impact may reflect its unique design philosophy: rather than relying solely on clinical guideline content, the platform’s architecture was explicitly mapped to patient discourse themes extracted through social media analysis [12]. This alignment between platform content and patient-identified concerns may have enhanced engagement, self-efficacy, and ultimately self-management capacity, reducing reliance on emergency care for symptom exacerbations.

The subgroup findings offer mechanistic insights. The stronger effects among younger adults (19–39 years) align with expected digital engagement patterns and suggest that mobile-first self-management tools may be particularly effective for this demographic. The disproportionate reduction in IBS-D–attributed ED visits is consistent with the prominent role of dietary management content within the platform, informed by the ‘diet and triggers’ theme that constituted 15.1% of IBS social media discourse [12]. Patients with IBS-D may be more responsive to dietary self-management tools because symptom exacerbations are often precipitated by identifiable dietary triggers amenable to the platform’s personalized tracking and avoidance algorithms. The weaker effects for IBS-C and IBS-unspecified may reflect the greater complexity of constipation management, which is less amenable to dietary self-management alone.

The dose-response relationship between platform registration rates and ED visit reductions (Table 5) strengthens the causal interpretation. Areas with higher registration rates (Q4: 3.14%) showed substantially larger level changes (−5.42 per 100,000) compared to lower-registration areas (Q1: −1.78, non-significant), with a significant trend of −1.87 per 1% registration increase (p = 0.008). This gradient argues against confounding by unmeasured area characteristics and supports a plausible mechanism through which individual-level platform engagement aggregates to population-level utilization changes.

**Table 5.** Dose-Response Relationship Between Platform Registration Rate and IBS-Attributed ED Visit Reduction.

| Platform Registration<br>Rate Quartile | Mean<br>Registration<br>(%) | Level Change<br>($\beta$) | 95% CI | p-value |
| --- | --- | --- | --- | --- |
| Q1 (lowest) | 0.84 | -1.78 | -3.94 to +0.38 | 0.107 |
| Q2 | 1.42 | -2.91 | -4.88 to -0.94 | 0.004 |
| Q3 | 2.08 | -3.87 | -6.02 to -1.72 | <0.001 |
| Q4 (highest) | 3.14 | -5.42 | -7.81 to -3.03 | <0.001 |
| Trend per 1% increase | — | -1.87 | -3.24 to -0.50 | 0.008 |

Note: Registration rate quartiles calculated across 16 sub-metropolitan districts within the four intervention areas. The dose-response trend was estimated via meta-regression of district-level CITS estimates on registration rates. CI = confidence interval.

Methodologically, the CITS design provides several advantages over uncontrolled ITS for evaluating digital health interventions [14,15]. By incorporating eight concurrent control areas, the design accounts for contemporaneous threats to internal validity — most notably the COVID-19 pandemic, which produced transient shifts in healthcare utilization across all areas. The null placebo date tests and the stability of estimates across pre-intervention window lengths support the validity of the parallel trends assumption. The corroboration of segmented regression findings by ARIMA transfer function models provides additional confidence in the temporal attribution of effects. These methodological considerations are particularly important given the challenges of evaluating population-level digital health interventions, where randomization is often infeasible and secular trends in healthcare utilization introduce significant confounding potential [16,17].

This study contributes to the growing evidence base on translating social media analytics into actionable public health interventions. While prior research has demonstrated the feasibility and value of applying NLP techniques — including sentiment analysis and topic modeling — to extract patient insights from platforms like X.com [12], evidence on downstream clinical and population-level impacts of social media–informed interventions has been limited. Our findings suggest that the systematic extraction of patient discourse themes can inform the design of digital health tools that are both clinically effective and aligned with patient priorities, potentially enhancing user engagement and real-world effectiveness. This is consistent with the emerging paradigm of patient-centered digital health design, which integrates user-generated data into intervention development to improve relevance and uptake [26,27].

### 4.1 Limitations

Several limitations merit consideration. First, the CITS design estimates population-level associations and cannot establish individual-level causal mechanisms; we cannot confirm that the observed reductions were mediated specifically by platform users changing their health-seeking behavior. Second, the intervention areas were not randomly assigned, and despite comparable baseline characteristics, residual confounding by unmeasured area-level factors remains possible. Third, the platform registration rate of 1.83% is relatively low, raising questions about how population-level effects were achieved; spillover effects through social networks, community health agent amplification, and herd-like information effects represent plausible explanations requiring further investigation. Fourth, the NHIS claims data do not capture clinical severity at the individual level, precluding analysis of whether ED visit reductions represent appropriate substitution of self-management for emergency care or potentially inappropriate avoidance of necessary care. Fifth, the social media discourse that informed the platform’s content was based on English-language posts [12], and cultural adaptation to the Korean context, while conducted by a multidisciplinary panel, may not fully capture Korean-language IBS discourse patterns. Sixth, generalizability to rural areas and older populations may be limited given the concentration of effects in urban metropolitan areas and younger age groups. Finally, the 42-month post-intervention period, while substantial, may be insufficient to assess long-term sustainability of effects.

### 4.2 Clinical and Policy Implications

These findings carry implications for health system policy in South Korea and beyond. The integration of the Jang Geongang platform within the existing NHIS chronic disease management framework demonstrates scalability within single-payer universal health systems. The estimated annual avoidance of 1,790 ED visits in the pilot areas, at an average NHIS reimbursement of □234,000 per IBS-attributed ED visit, suggests potential annual savings of approximately □419 million (USD 320,000) in direct ED costs alone, not accounting for downstream hospitalization reductions and indirect costs. The dose-response findings suggest that investment in platform promotion and enrollment could yield proportional returns. More broadly, the study supports the emerging model of using social media analytics as a formative research tool for public health intervention design [12], applicable to other chronic conditions where patient-generated discourse reveals unmet needs. South Korea’s NHIS, with its comprehensive claims infrastructure, is well-positioned to expand this approach to additional chronic conditions including inflammatory bowel disease, gastroesophageal reflux disease, and chronic liver disease.

## 5. Conclusions

A digital self-management platform for IBS, designed using social media–derived patient discourse insights from NLP-based topic modeling and sentiment analysis, was associated with sustained population-level reductions in IBS-attributed emergency department utilization when evaluated using a controlled interrupted time series design with 84 monthly observations across South Korean metropolitan areas. The estimated 23.8% cumulative relative reduction in IBS-attributed ED visits, corroborated by ARIMA transfer function models and a dose-response gradient, provides robust evidence for the public health impact of translating social media analytics into scalable digital health interventions for functional gastrointestinal disorders.

## Data Availability

All data produced in the present work are contained in the manuscript

## References

1. Han SH, Lee OY, Bae SC, et al. Prevalence of irritable bowel syndrome in Korea: population-based survey using the Rome II criteria. J Gastroenterol Hepatol. 2006;21(11):1687–1692.

2. Park DW, Lee OY, Shim SG, et al. The differences in prevalence and sociodemographic characteristics of irritable bowel syndrome according to Rome II and Rome III. J Neurogastroenterol Motil. 2010;16(2):186–193.

3. Kim YS, Kim N. Functional dyspepsia and irritable bowel syndrome in Korea: prevalence and impact on quality of life. J Neurogastroenterol Motil. 2018;24(2):182–196.

4. Shin SY, Cha BK, Kim WS, et al. The burden of irritable bowel syndrome in Korea: analysis using national health insurance data. Korean J Gastroenterol. 2020;75(6):316–323.

5. Jung HK, Kim YH, Park JY, et al. Estimating the burden of irritable bowel syndrome: analysis of a nationwide Korean database. J Neurogastroenterol Motil. 2014;20(2):242–252.

6. Ford AC, Sperber AD, Corsetti M, Camilleri M. Irritable bowel syndrome. Lancet. 2020;396(10263):1675–1688.

7. Heitkemper M, Jarrett M, Jun SE. Update on irritable bowel syndrome programme of research. J Adv Nurs. 2013;69(5):1025–1034.

8. Murray E, Burns J, See TS, et al. Interactive health communication applications for people with chronic disease. Cochrane Database Syst Rev. 2005;(4):CD004274.

9. Drossman DA, Tack J. Rome Foundation clinical diagnostic criteria for disorders of gut-brain interaction. Gastroenterology. 2022;162(3):675–679.

10. Sinnenberg L, Buttenheim AM, Padrez K, et al. Twitter as a tool for health research: a systematic review. Am J Public Health. 2017;107(1):e1–e8.

11. Eysenbach G. Infodemiology and infoveillance: framework for an emerging set of public health informatics methods to analyze search, communication and publication behavior on the internet. J Med Internet Res. 2009;11(1):e11.

12. Shankar R, Yip A. Sentiment analysis and topic modeling of social media data to explore public discourse on irritable bowel syndrome. Sci Rep. 2025;15(1):10829. doi:10.1038/s41598-025-08599-7.

13. Wagner AK, Soumerai SB, Zhang F, Ross-Degnan D. Segmented regression analysis of interrupted time-series studies in medication use research. J Clin Pharm Ther. 2002;27(4):299–309.

14. Bernal JL, Cummins S, Gasparrini A. Interrupted time series regression for the evaluation of public health interventions: a tutorial. Int J Epidemiol. 2017;46(1):348–355.

15. Lopez Bernal J, Cummins S, Gasparrini A. The use of controls in interrupted time series studies of public health interventions. Int J Epidemiol. 2018;47(6):2082–2093.

16. Kontopantelis E, Doran T, Springate DA, Buchan I, Reeves D. Regression based quasi-experimental approach when randomisation is not an option: interrupted time series analysis. BMJ. 2015;350:h2750.

17. Cochrane Effective Practice and Organisation of Care (EPOC). EPOC resources for review authors: interrupted time series (ITS) analyses. 2017. Available at: epoc.cochrane.org.

18. Kwon S. Thirty years of national health insurance in South Korea: lessons for achieving universal health care coverage. Health Policy Plan. 2009;24(1):63–71.

19. Bhaskaran K, Gasparrini A, Hajat S, Smeeth L, Armstrong B. Time series regression studies in environmental epidemiology. Int J Epidemiol. 2013;42(4):1187–1195.

20. Newey WK, West KD. A simple, positive semi-definite, heteroskedasticity and autocorrelation consistent covariance matrix. Econometrica. 1987;55(3):703–708.

21. Prais SJ, Winsten CB. Trend Estimators and Serial Correlation. Cowles Commission Discussion Paper No. 383. Chicago; 1954.

22. Box GEP, Tiao GC. Intervention analysis with applications to economic and environmental problems. J Am Stat Assoc. 1975;70(349):70–79.

23. Hyndman RJ, Khandakar Y. Automatic time series forecasting: the forecast package for R. J Stat Softw. 2008;27(3):1–22.

24. Pinheiro J, Bates D, R Core Team. nlme: linear and nonlinear mixed effects models. R package version 3. 1–164. 2023.

25. Lalouni M, Ljotsson B, Bonnert M, et al. Internet-delivered cognitive behavioral therapy for children with pain-related functional gastrointestinal disorders: feasibility study. JMIR Ment Health. 2017;4(3):e32.

26. Wicks P, Massagli M, Frost J, et al. Sharing health data for better outcomes on PatientsLikeMe. J Med Internet Res. 2010;12(2):e19.

27. Moorhead SA, Hazlett DE, Harrison L, Carroll JK, Irwin A, Hoving C. A new dimension of health care: systematic review of the uses, benefits, and limitations of social media for health communication. J Med Internet Res. 2013;15(4):e85.

28. Cruz Rivera S, Liu X, Chan AW, et al. Guidelines for clinical trial protocols for interventions involving artificial intelligence: the SPIRIT-AI extension. Lancet Digit Health. 2020;2(10):e549–e560.

29. Penfold RB, Zhang F. Use of interrupted time series analysis in evaluating health care quality improvements. Acad Pediatr. 2013;13(6 Suppl):S38–S44.

30. Jandoc R, Burden AM, Mamdani M, Lévesque LE, Cadarette SM. Interrupted time series analysis in drug utilization research is increasing: systematic review and recommendations. J Clin Epidemiol. 2015;68(8):950–956.

